# Cardiopulmonary Exercise Testing is Safe in Children and Adolescents with Cardiomyopathy

**DOI:** 10.1101/2025.05.20.25328041

**Authors:** Kelsey Iguidbashian, Julie Fernie, Kaitlin Olson, Jean Cavanaugh, Shelley D. Miyamoto, Roni Jacobsen

## Abstract

**Background:** Cardiopulmonary exercise testing (CPET) has been recommended in pediatric patients with hypertrophic cardiomyopathy (HCM); however, safety has not been well defined. Studies in adults with cardiomyopathy (CM) consistently show reduced maximal and submaximal exercise capacity and impaired ventilatory efficiency compared to matched controls, with mixed data in children. This single-center retrospective study aimed to identify differences in adverse events and exercise parameters in children and adolescents with CM versus matched controls.

**Methods:** From 2014 to 2023, 227 CPETs were completed in 141 patients with CM, ages 6 to 22 years. Only maximal CPETs were included for control-matched sub analysis, resulting in 113 tests in 83 CM patients. Subjects were matched with healthy controls based on sex, age, body mass index (BMI), body surface area (BSA), and stress exercise modality to evaluate for differences in heart rate (HR), systolic blood pressure (SBP), electrocardiogram (ECG) changes, incidence of adverse events (ischemia, SBP drop, and arrhythmias), exercise-induced symptoms, and peak exercise parameters.

**Results:** There were 4 CM tests (all HCM, 1.8%) terminated due to adverse events: SBP drop (2) and non-sustained ventricular tachycardia (2). Control patients reported significantly more symptoms of chest pain and dizziness with exercise. Maximal and submaximal oxygen consumption (VO_2_), SBP, HR, and minute ventilation (VE) were lower in CM patients.

**Conclusion:** In our cohort, CPET testing in children and adolescents with CM was safe. Maximal and submaximal exercise capacity was reduced in CM patients, likely due to reduced cardiac output, with lower HR and VE at peak exercise. Future studies may identify if exercise parameters can be used to risk stratify patients or predict outcomes.

## Introduction

Pediatric cardiomyopathies (CMs) are rare, affecting roughly 1.1 to 1.5 per 100,000 individuals (1). Phenotypic presentations vary greatly with outcomes, ranging from normal cardiac function to transplantation and death. Thus, there have been efforts to stratify patients to better understand long term risks, including prognostication with CPET.

Cardiopulmonary exercise tests (CPETs) are a tool that have been used in limited capacity to risk stratify in the setting of CM. CPETs have been shown to generally be a safe procedure in adults with high-risk cardiac diseases (including HCM) (2–4) and pediatric and adolescent patients with pulmonary hypertension (5). Historically, pediatric and adolescent patients with hypertrophic CM (HCM), restrictive CM (RCM), and dilated CM (DCM) have been considered higher risk for exercise testing, especially in the presence of congestive heart failure and arrhythmias (6). There is minimal literature on the safety of CPETs in pediatric and adolescent CM populations. CPET has been recommended in adults with HCM, with recent studies describing use in pediatric HCM patients (7–10). Additionally, peak oxygen consumption (VO_2_ peak) has been used as a prognosticator for outcomes in pediatric HCM, DCM, and RCM (8, 9, 11–14). With limited data related to safety given concern that these patients may be high risk for adverse events, CPET in children and adolescents with CM is often avoided (6).

Research shows the utility of CPET for functional assessment and prognostic measurement in pediatric and adult patients with HCM, but there is very limited research on CPET in children and adolescents with other types of CM. Patients with HCM generally have impaired cardiovascular fitness with lower peak VO_2_ and peak power (8–10), likely due to an inability to augment cardiac output due to reduction in stroke volume and chronotropic reserve (15). During CPET, ectopy and lower percent-predicted peak VO_2_ have been associated with worse outcomes (heart failure, sudden cardiac death) (8, 9). Given the lack of data regarding CPET data in children and adolescents with CM, this study aimed to: 1) assess safety of CPET in patients with CM, 2) identify differences in CPET parameters for children and adolescents with CM versus healthy children and adolescents and 3) assess changes in CPET parameters over serial tests in children and adolescents with CM.

## Methods

The Colorado Multiple Institutional Review Board approved the protocol with a waiver of consent in view of the retrospective nature of the study. All procedures performed were part of routine clinical care. This single-center retrospective study analyzed all CPETs completed in patients with phenotypic (+/- genotype positive) CM (HCM, DCM, RCM, left ventricular non-compaction (LVNC), arrhythmogenic right ventricular cardiomyopathy (ARVC), and other) between 2014 and 2023. There were 227 CPETs performed in 141 CM patients, aged 6 to 22 years.

All CPETs were completed on an MGC Diagnostics Ultima CPX metabolic cart (MGC Diagnostics Saint Paul, MN, USA) that was maintained and calibrated per manufacturer guidelines. Patients completed a pulmonary function test (PFT) prior to exercise. The standard exercise modality in our lab is the cycle ergometer (Lode Corival CPET, Gronigen, Netherlands) with a ramp protocol of approximately 3 watts/kg starting at zero watts for a targeted maximal exercise duration of 8 to 12 minutes. Tests were performed on the treadmill using the Bruce protocol in children and adolescents who were not tall enough for the cycle ergometer. Patients underwent continuous metabolic monitoring during exercise, including oxygen consumption (VO_2_), carbon dioxide production (VCO_2_), minute ventilation (VE), and respiratory exchange ratio (RER). Heart rate (HR) and rhythm were measured by ECG. Cardiopulmonary exercise values were reported as 10-second averages. Peak VO_2_, peak heart rate, peak VE, and peak RER were defined as the highest values for each variable during exercise. Submaximal VO_2_ was identified using the V-slope method. Oxygen saturation (SpO_2_) was measured with a forehead probe and pulse oximeter (Nellcor PM1000N, Medtronic, Minneapolis, MD). Oxygen (O_2_) pulse was calculated as peak VO_2_ (mL/min) divided by peak HR. Blood pressure was measured by auscultation using an appropriately sized cuff. A SunTech Tango M2 system (Morrisville, NC) was used to auto-inflate the blood pressure cuff, and blood pressure was auscultated by the supervising exercise physiologist. Blood pressure was measured at rest, every 2 minutes during exercise and recovery, and at peak exercise (defined as within 30 seconds of peak exercise).

Patients were asked about any exercise-related symptoms (chest pain, shortness of breath, lightheadedness, palpitations, headache) every 2 minutes throughout exercise and recovery and were instructed to alert the supervising exercise physiologist about any symptoms that developed in the interim. Peak workload on the cycle ergometer was identified as the peak watts achieved.

Only maximal CPETs, defined per Takken et al. as an RER ≥ 1.0 and/or subjective assessment by the supervising exercise physiologist (16), were included for matched comparison. There were 113 maximal CPETs in 83 unique CM patients that were included in the matched comparison. CM subjects were matched with controls based on sex, age, height, weight, body mass index (BMI), body surface area (BSA), and stress exercise modality (cycle ergometer versus treadmill). Control patients were referred for CPET for symptoms with exercise and/or family history of cardiac disease. Participants were classified as controls if they had a history of a normal echocardiogram, or if no echocardiogram had been completed, if they had a normal cardiac examination and electrocardiogram. Children and adolescents with structural abnormalities or channelopathies such as long QT syndrome or catecholaminergic polymorphic ventricular tachycardia were excluded from the control cohort. Evaluation included baseline PFT; baseline and peak HR; baseline and peak blood pressure (BP); ECG changes; incidence of adverse events, including ischemia (ST segment changes), decrease in systolic blood pressure from baseline, and arrhythmias (defined as 3 or more subsequent beats of atrial or ventricular ectopy); and exercise-induced symptoms between groups. Exercise parameters including VO_2_ peak, submaximal VO_2_ (VO_2_ at ventilatory anaerobic threshold), peak VE, peak watts, VE/VCO_2_ slope at anaerobic threshold, peak HR, O_2_ pulse, peak respiratory rate, and VO_2_/work rate were also compared between groups. Statistical significance was set at p= 0.05.

For patients that underwent serial CPETs (2 or more), echocardiographic, electrocardiographic, Holter monitor, and laboratory data were included for analysis if completed within 1 month of the most recent CPET. Echocardiogram variables included left ventricular (LV) mass (g/m^2^), maximal intraventricular septal diameter in diastole (mm), LV outflow tract maximal velocity (m/s), and qualitative right ventricular and LV systolic function. Laboratory data included BNP, NT-pro BNP, creatinine, and cystatin-C. Documentation of adjustments in medical or diuretic therapy (increased, decreased, or stable) was also recorded from chart review.

### Variable manipulation

To create the “Cardiomyopathy type” variable, patients diagnosed with HCM, DCM, RCM, and LVNC were separated into distinct categories. All other classifications for CM patients (ARVC and Mixed) were collapsed into an “Other” category due to small sample size. The “Atrial ECG findings” variable was defined as an atrial pair and/or atrial run (3 or more beats) at rest or during exercise. “Ventricular ECG findings” was defined as multiple premature ventricular contractions (PVCs), ventricular pair, and/or a ventricular run (3 or more beats) at rest or during exercise.

### Statistical analysis

For the matched datasets, the first recorded data point was used and then further stratified by patients who completed the CPET on the cycle ergometer versus the treadmill. In total, 69 CM CPETs were matched against 69 control CPETs on the cycle ergometer, while 14 CM CPETs were matched to 14 control CPETs on the treadmill.

Many of the numeric variables displayed skew. Therefore, the median (Q1, Q3) was used to represent the data descriptively and the Wilcoxon Rank Sum test was used to calculate p-values. Categorical variables were summarized by N (%), and p-values were calculated using the Fisher’s exact test to account for small sample sizes.

For serial testing, demographic, exercise, and clinical data was summarized using median (interquartile range) for numeric variables and frequency and percentage for categorical variables. For each of the exercise and clinical variables in the study, we calculated the change between a patient’s first and last visit. Then, due to the variability in number of patient visits and time between each visit, we divided the change by time (years) between visits. This resulted in amount of change per year for each patient. Pearson’s correlation coefficient was used to identify exercise and clinical variables that were related to each of the outcome variables. Variables with p < 0.20 were considered for multivariate linear regression and backward selection was used to remove non-significant variables to determine the final model. Significance level was set to an p= 0.05 level. R version 4.2.3 for MacOS was used for all statistical analysis.

## Results

Of the 141 CM patients, the majority had DCM (64), followed by HCM (56), LVNC (12), Other (5), and RCM (4). Maximal CPET was achieved in 100% of RCM patients, 67% of LVNC patients, 64% of HCM patients, 60% of Other CM, and 50% of DCM. Sixty-nine patients (31 HCM, 23 DCM, 4 RCM, 8 LVNC, and 3 others) completed CPET on the cycle ergometer with a median age of 14 years. Fourteen patients completed CPET on the treadmill (8 DCM, 5 HCM, 1 LVNC) with a median age of 9 years. Of the 227 CPETs performed, 4 separate patients (all HCM, 1.8% of total tests) were terminated due to adverse events: 2 due to drop in SBP and 2 due to non-sustained ventricular tachycardia.

Of the 69 patients who underwent cycle ergometer testing, there was no difference in sex, age, race/ethnicity, height, weight, BMI, or BSA between CM and controls (Table 1). There was no difference in baseline HR, but the peak HR was significantly lower (181 bpm versus 189 bpm, p< 0.001) in CM patients (Table 2). Eighteen (26%) CM patients on a beta blocker had significantly lower peak exercise HR compared to CM patients not on a beta blocker (169.9 bpm versus 181.2 bpm, p=0.01, Table 3). There was no difference in baseline SBP, although peak SBP was significantly lower in CM patients compared to controls (143 mmHg versus 154 mmHg, p= 0.021, Table 2.) There was no significant difference in baseline or peak exercise SBP for CM patients on or off an ACE (angiotensin-converting enzyme) inhibitor (p=0.333) or beta blocker (p=0.271, Table 4). There was inadequate power to evaluate differences between CM and controls on CPETs completed on the treadmill due to small sample size

**Table 1.**
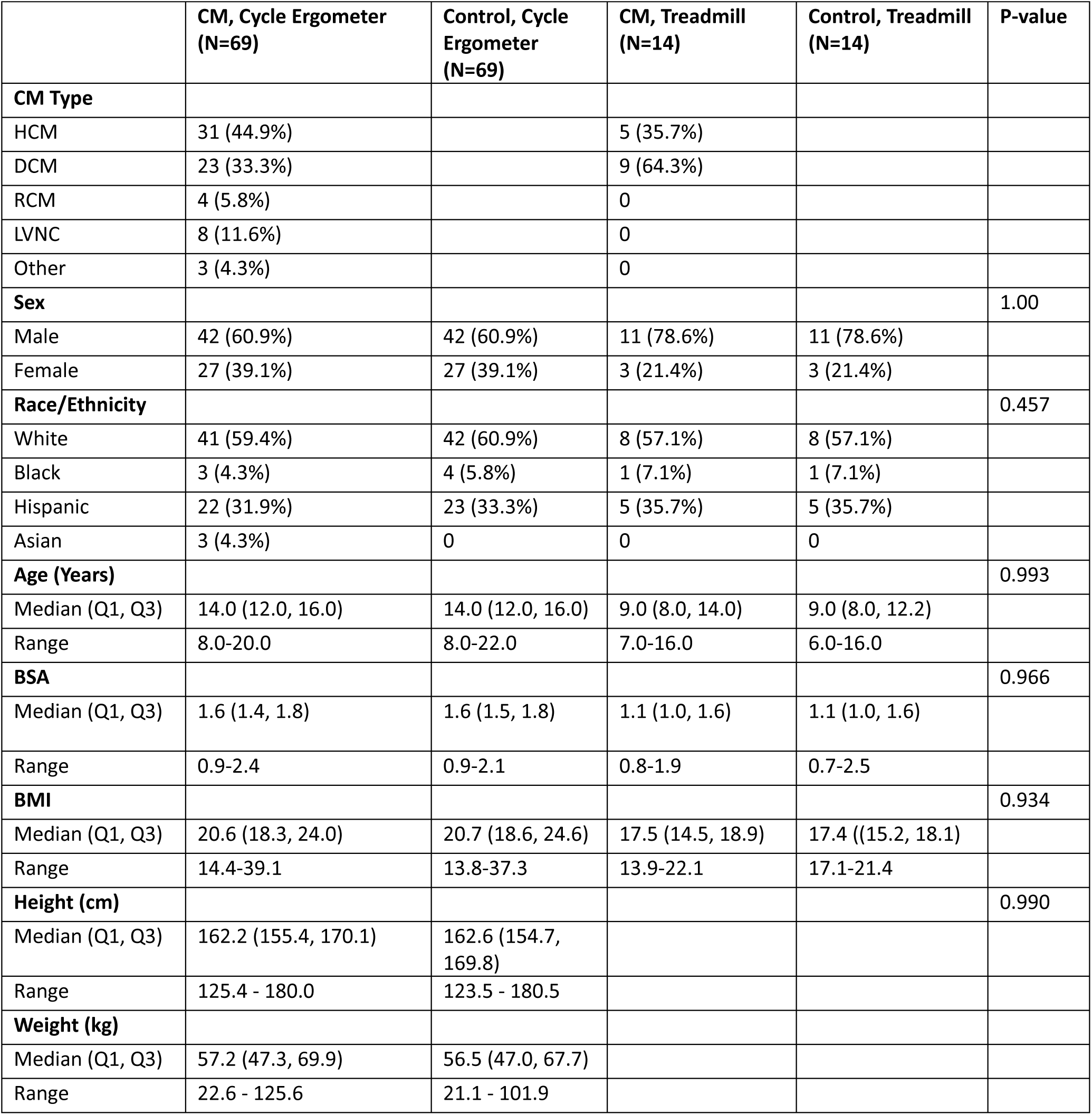
Baseline demographics for CM and control patients undergoing cycle ergometer and treadmill CPET.

**Table 2.**
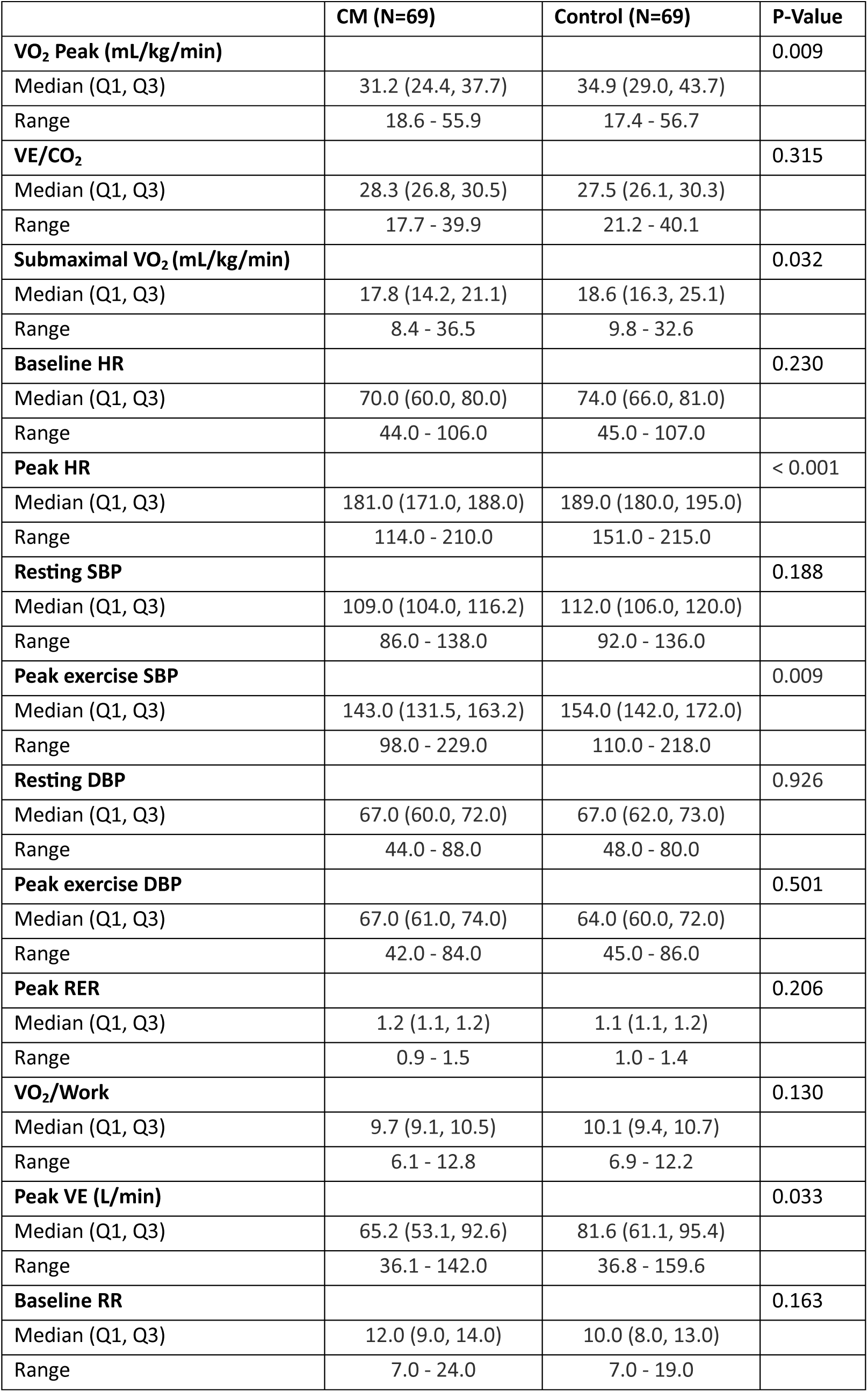
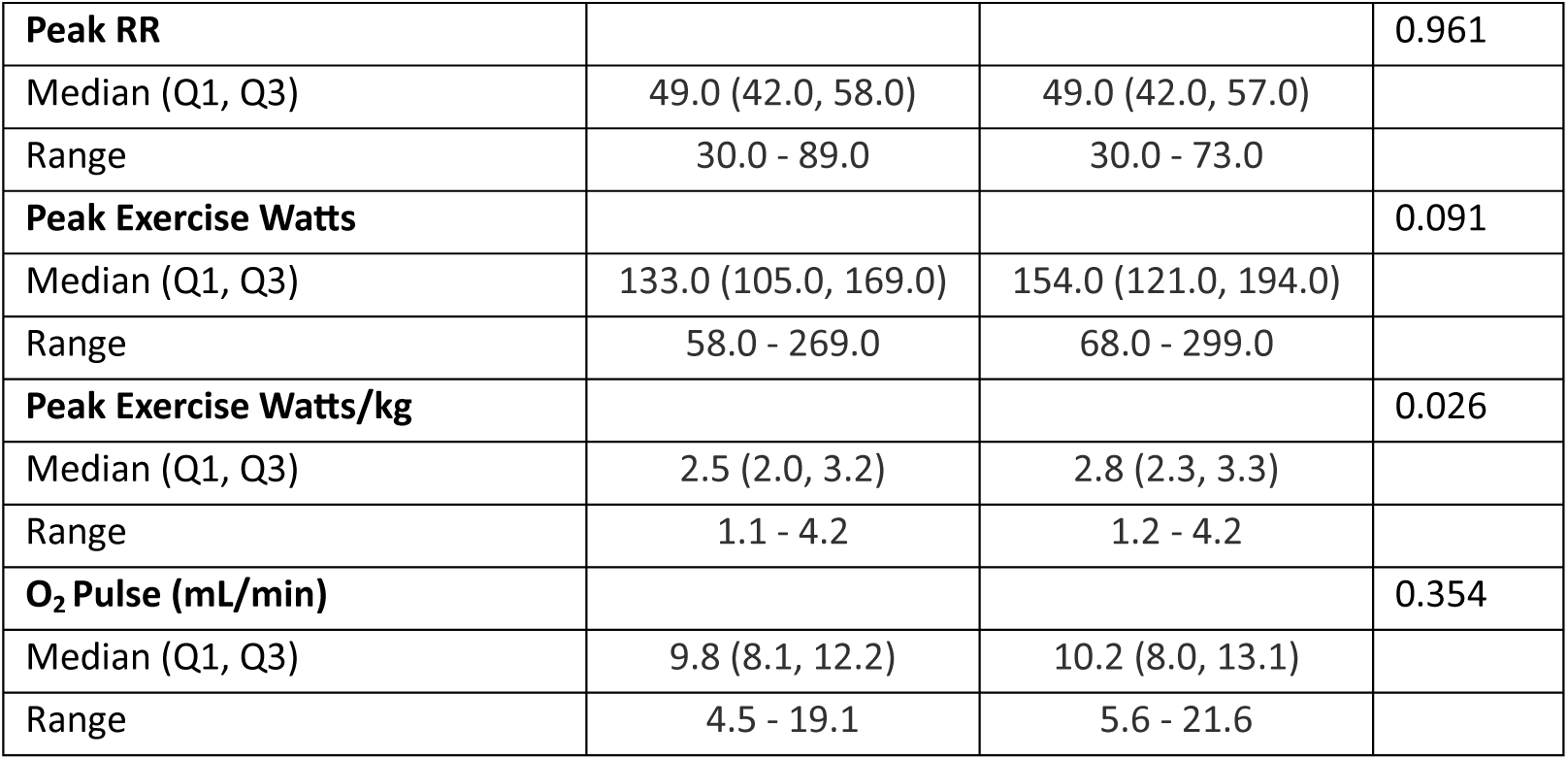
Exercise variables for CM and control patients undergoing cycle ergometer CPET.

**Table 3.**
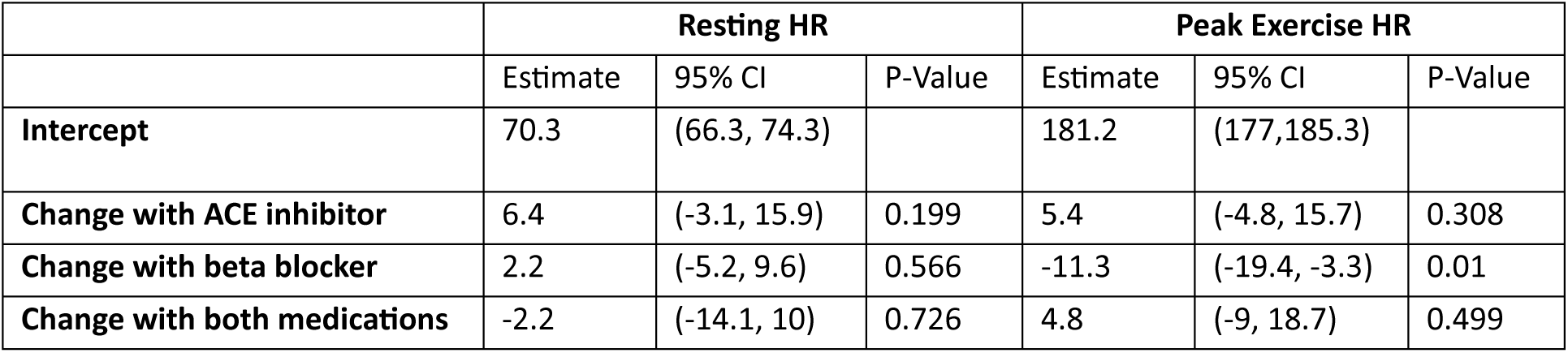
Baseline and exercise heart rate in CM patients on medical therapy.

**Table 4.** Baseline and exercise blood pressure in CM patients on medical therapy.

|  | Resting HR |  |  | Peak Exercise HR |  |  |
| --- | --- | --- | --- | --- | --- | --- |
|  | Estimate | 95% CI | P-Value | Estimate | 95% CI | P-Value |
| **Intercept** | 112.8 | (109.7, 115.9) |  | 157 | (150, 163.9) |  |
| **Change with ACE inhibitor** | -2 | (-9.6, 5.6) | 0.611 | -8.4 | (-25.2, 8.3) | 0.333 |
| **Change with beta blocker** | -1.4 | (-7.4, 4.5) | 0.646 | -7.5 | (-20.7, 5.6) | 0.271 |
| **Change with both medications** | -1.4 | (-11.4, 8.6) | 0.784 | -0.3 | (-22.3, 21.8) | 0.981 |

Symptoms during exercise testing varied between the CM and control groups (Table 5). Control patients reported significantly more symptoms of chest pain (p=0.007) and dizziness (p=0.004), while CM patients endorsed more headaches (p=0.042). There was no significant difference in symptoms of palpitations, nausea, lightheadedness, or shortness of breath between groups. Ventricular ectopy (defined as premature ventricular contractions, ventricular pair, and/or a ventricular run (3 or more beats)) occurred more frequently at baseline, mid-exercise, and during recovery in CM patients (Table 6). Two CM patients (1 RCM, 1 HCM) demonstrated ST depression at peak exercise compared to zero control patients, which was not statistically significant (p=0.154).

**Table 5.**
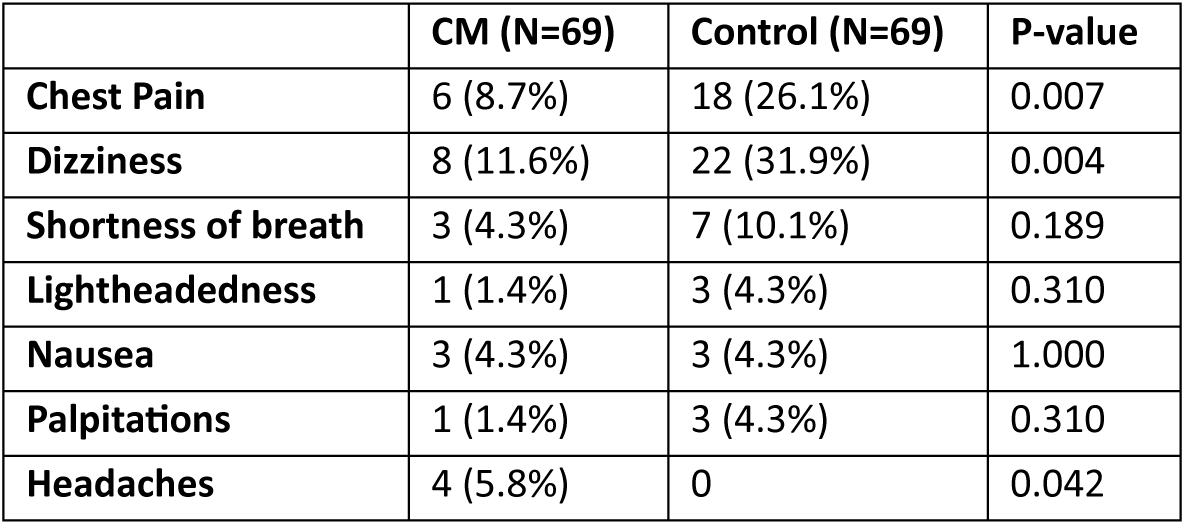
Reported symptoms in patients with CM versus controls with cycle ergometer CPET.

**Table 6.**
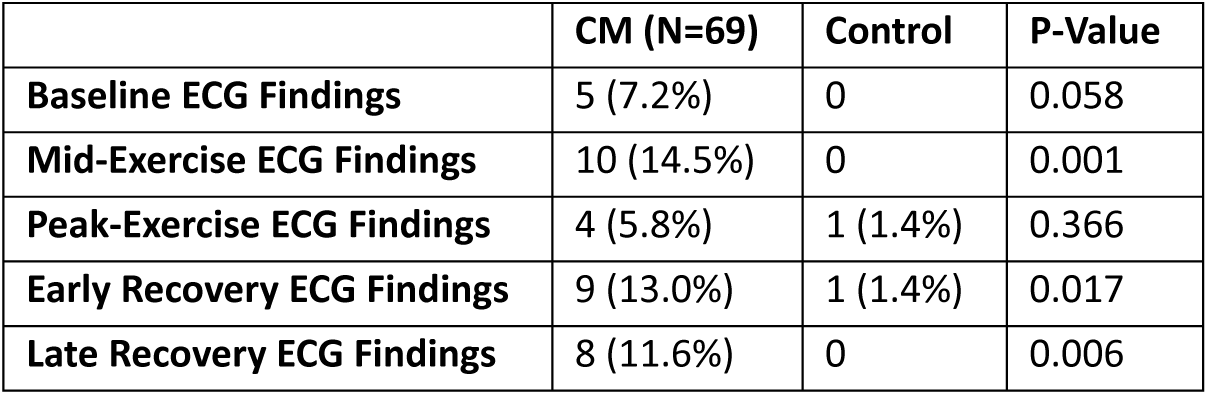
ECG findings in patients undergoing cycle ergometer CPET compared to controls.

Exercise parameters differed for patients with CM compared to healthy controls (Table 2). For patients tested on the cycle ergometer, VO_2_ peak (31.2 mL/kg/min versus 34.9 mL/kg/min, p = 0.009) and submaximal VO_2_ (17.8 versus 18.6 mL/kg/min, p = 0.032) was lower in CM patients. VE/VCO_2_ slope (28.3 versus 27.5, p = 0.315), VO_2_/work (9.7 mL/min/watt versus 10.1 mL/min/watt, p = 0.130), and O_2_ pulse (9.8 mL/beat versus 10.2 mL/beat, p = 0.354) was not different between CM and controls. O_2_ pulse was not altered by beta blocker use (p = 0.422) in CM. VE was lower in CM than controls (65.2 L/min versus 81.6 L/min, p = 0.033), despite no difference in peak respiratory rate between groups (49 breaths/minute, p = 0.961). For patients with maximal CPETs performed on the cycle ergometer, there was a higher prevalence of restrictive lung disease in CM versus controls (14.5% versus 2.2%, p=0.005). Restrictive lung disease correlated with lower VO_2_ peak (24.4 mL/kg/min versus 34.0 mL/kg/min, p = 0.001) compared to those with normal PFTs, while obstructive lung disease did not. Although not significantly different, patients with restrictive lung disease had lower VE (84.2 mL/min versus 80.6 mL/min, p=0.067) compared to patients with normal PFTs. There was no significant difference in the prevalence of restrictive or obstructive lung disease in CM versus control patients tested on treadmill.

Thirty CM patients underwent 2 or more serial CPETs on cycle ergometer within the identified time frame, 22 males (73%) and 8 females (27%) (Table 7). The median age for all serial testing was 13.5 years. Most serially tested patients had HCM (17), followed by DCM (8), LVNC (4), RCM (1), and Other (1). When comparing patient’s first and last test (change per years), there was a significant increase in peak work (watts), peak RER, O_2_ pulse, and peak VE and a significant decrease in peak VO_2_ (mL/kg/min), baseline HR, and baseline RR (Table 8).

**Table 7.**
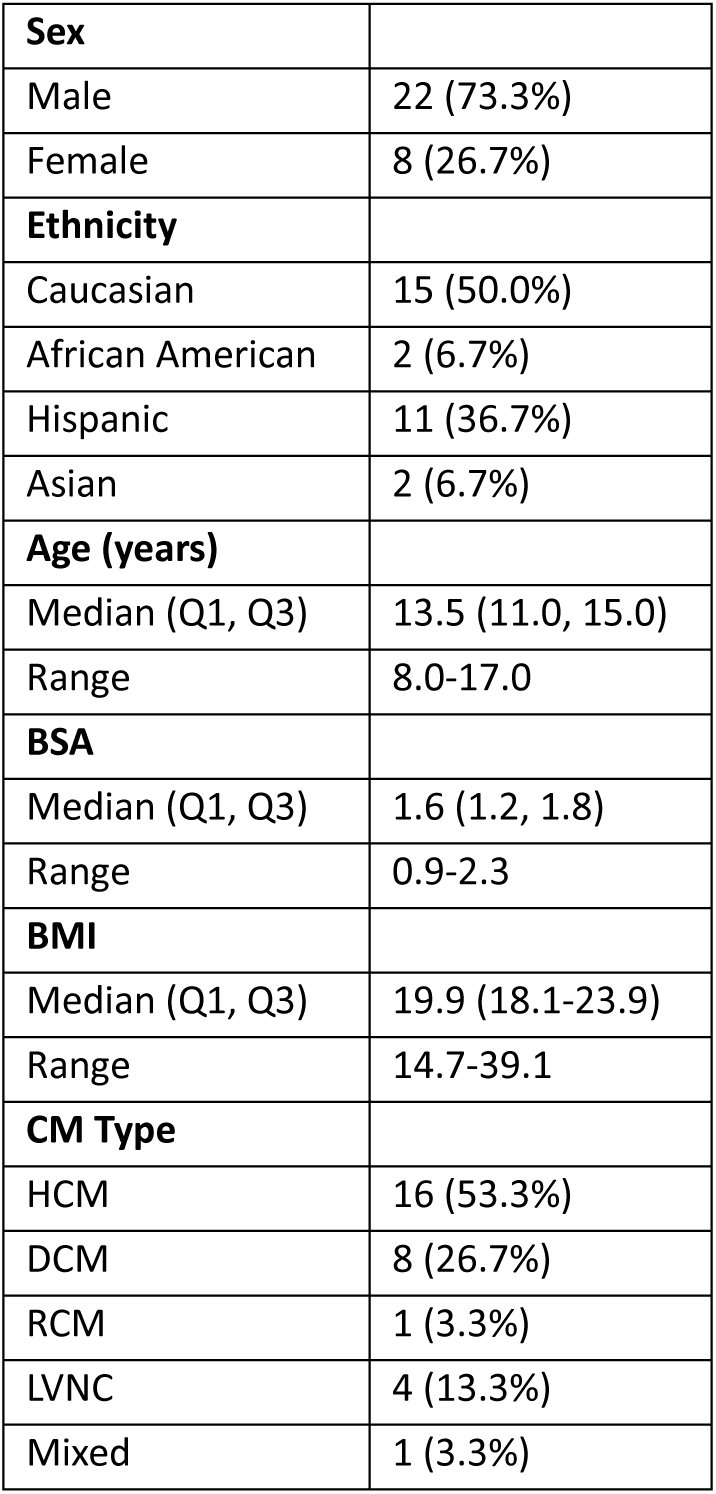
Summary statistics of baseline demographics of patients with serial CPETs (>2) on cycle ergometer.

**Table 8.**
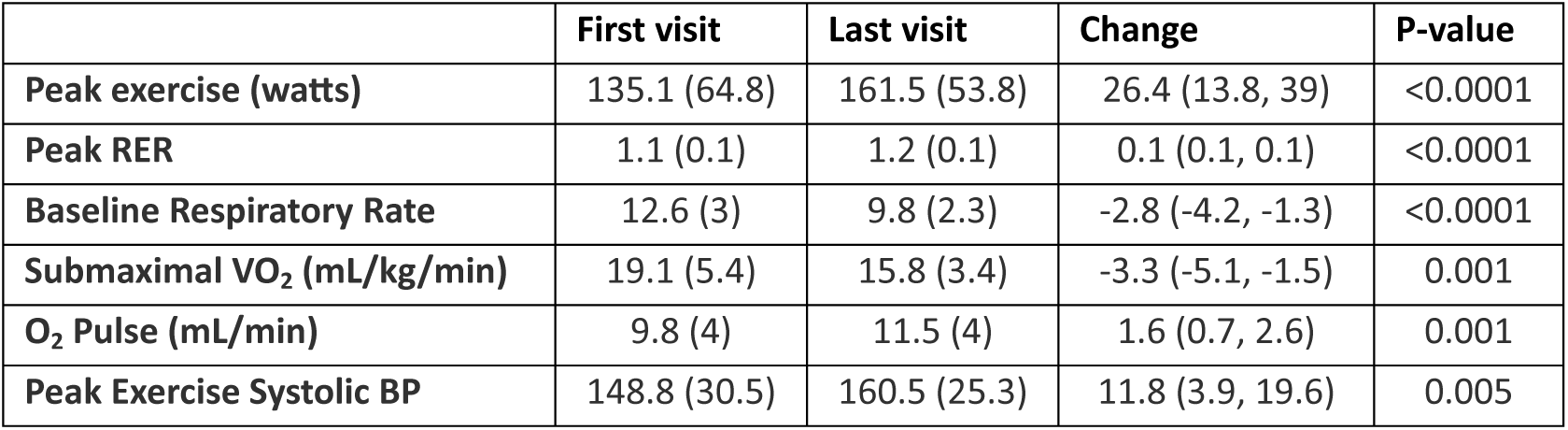
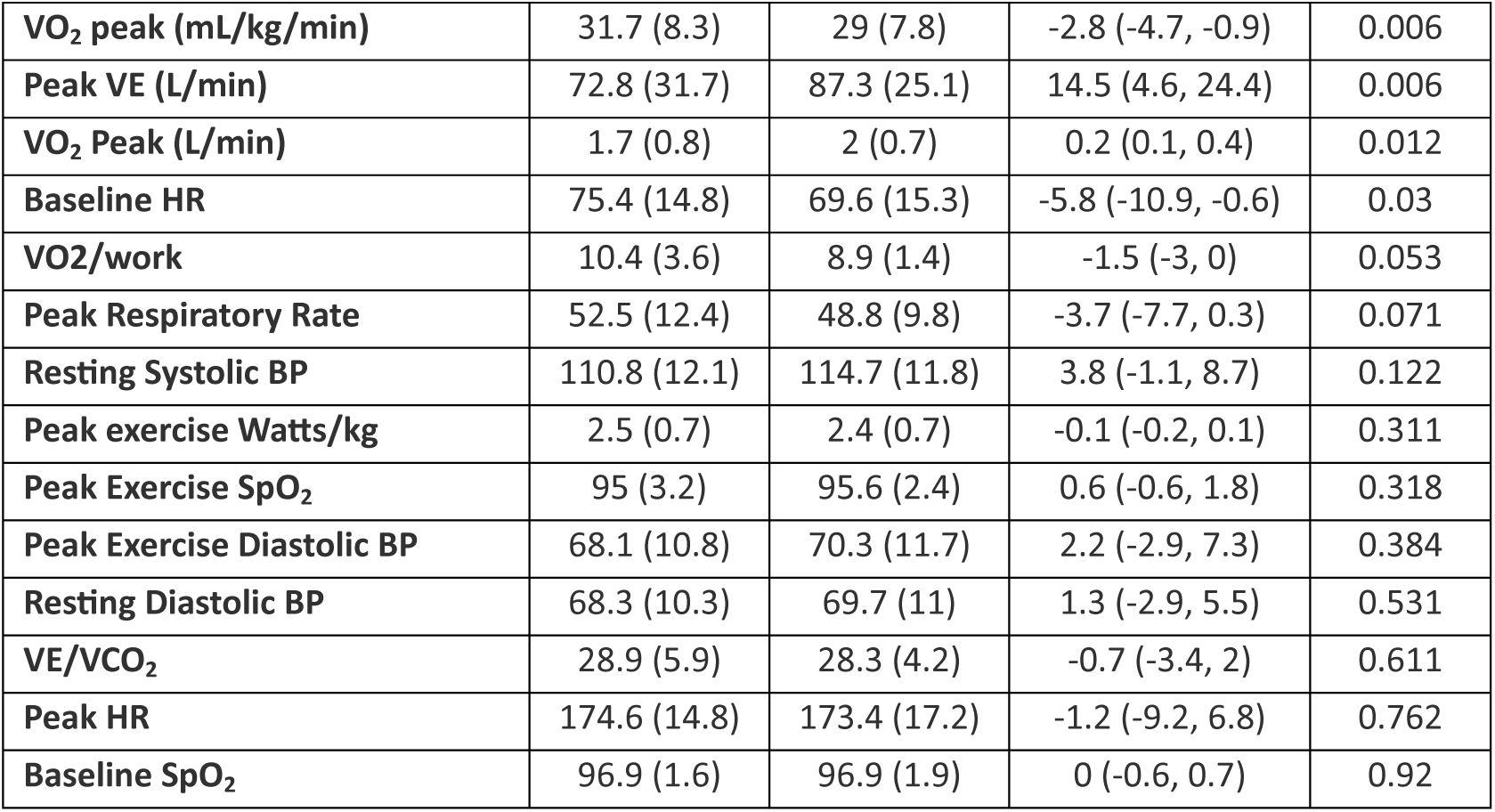
Changes in exercise variables between serial cycle ergometer CPETs.

Of patients with serial CPETs performed, 6 patients experienced heart transplant or death (3 HCM, 2 DCM, 1 RCM) during the study period. The median time between last CPET and transplant was 7 months (mean = 17.4 months), and the median time of CPET to death was 39.5 months (mean = 39.5 months). Ten patients without serial CPETs experienced heart transplant or death. The median time between CPET and transplant was 14 months (mean = 17 months), and the median time of CPET to death was 30 months (mean = 29 months). There were no significant differences in baseline BSA, resting SBP, or peak exercise SBP data for patients with serial testing that did not experience transplant or death compared to those that did (Table 9). There was no significant difference in change in serial demographic, exercise, or echocardiographic data (change per years) between groups. Due to absence of complete data, we were unable to evaluate a relationship between changes in exercise parameters and echocardiogram, ECG, Holter, and laboratory data. Minimal events (6 deaths and/or transplantation) prevented the ability to perform logistic regression to predict odds of transplant and/or death based on exercise data.

**Table 9.**
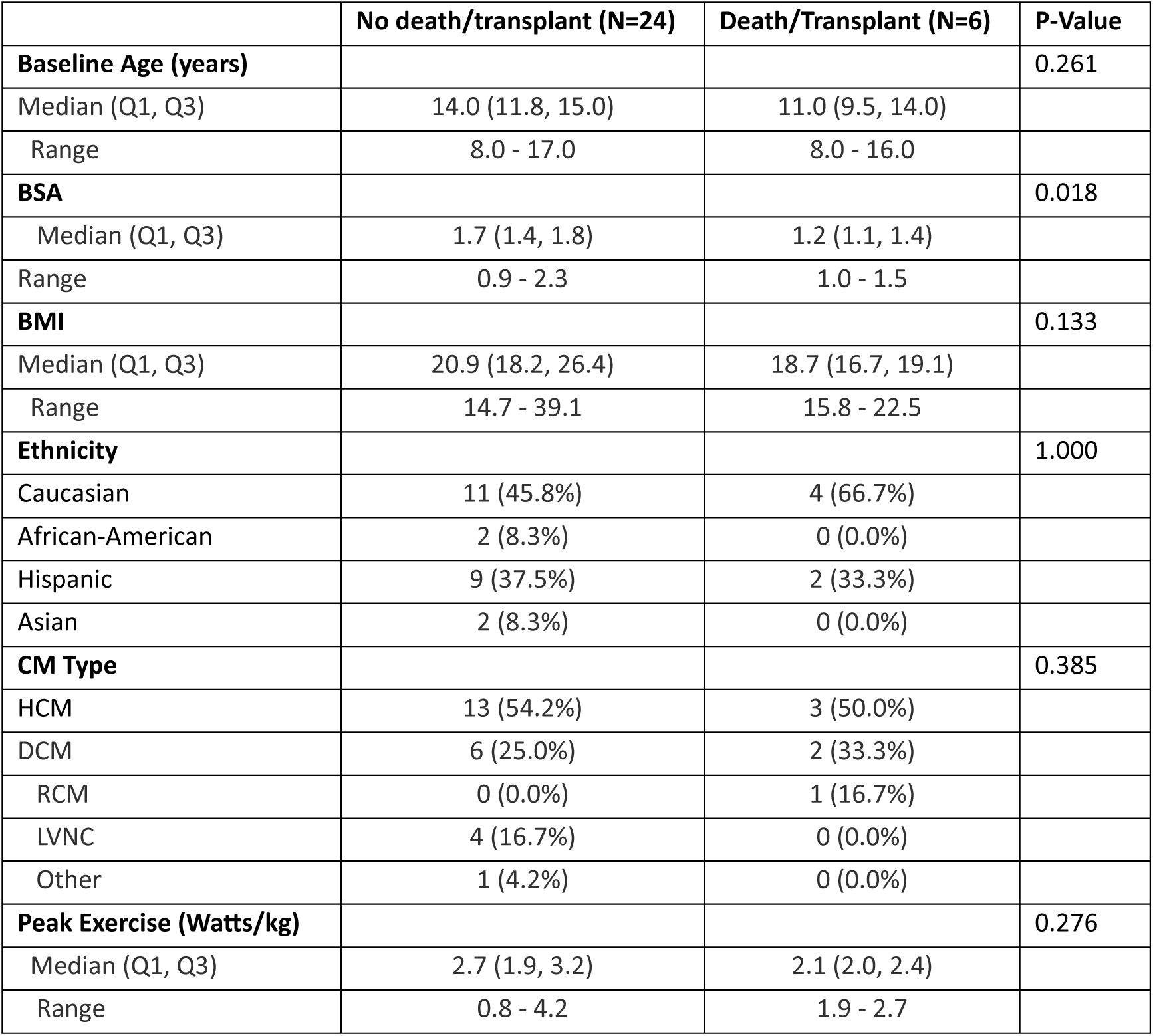
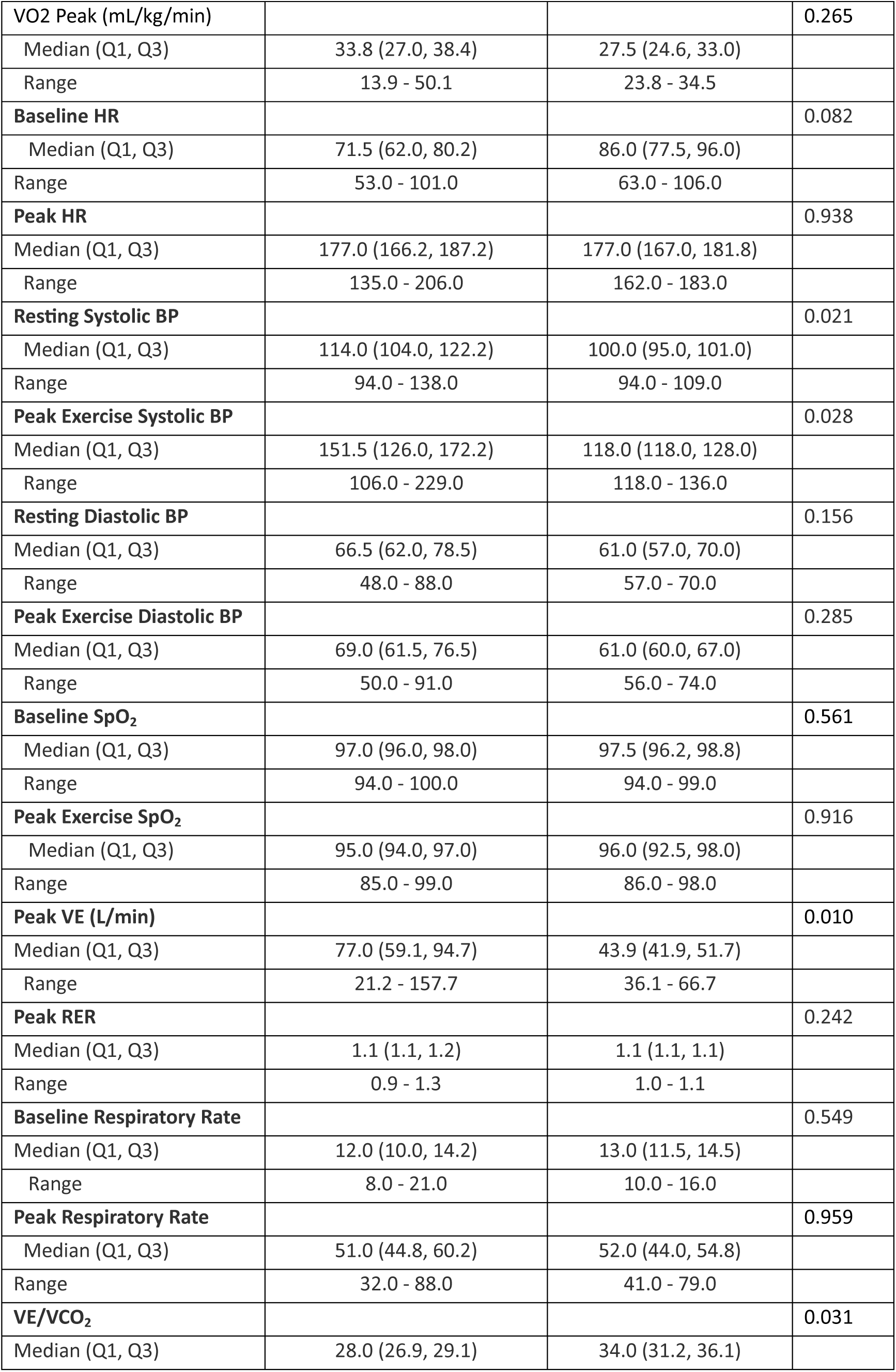
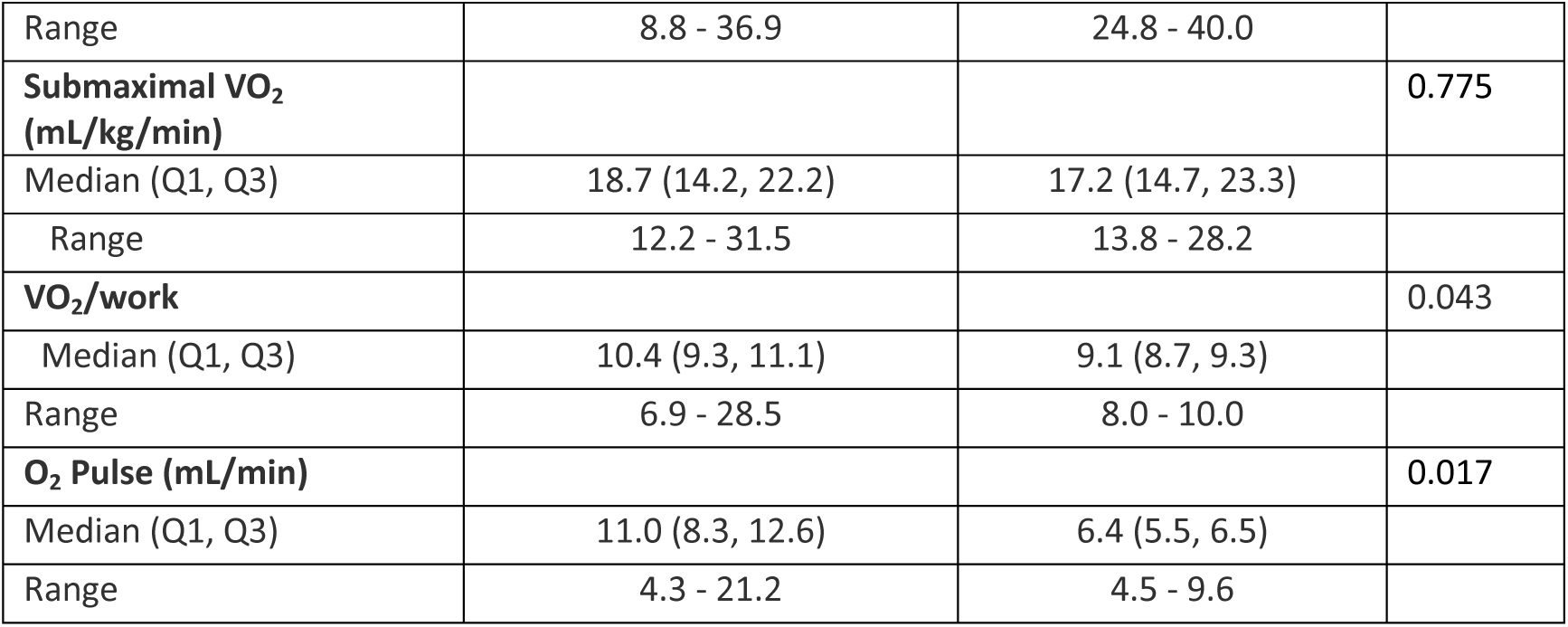
Baseline demographics, serial exercise variables, and echocardiogram variables in CM patients with events (heart transplant and/or death)

## Discussion

The primary goal of this study was to establish the safety of CPET in pediatric and adolescent patients with CM. Of the 227 tests performed in 141 patients with CM, 4 were terminated for adverse events (1.8%). Two tests were terminated for a drop in SBP and 2 for non-sustained ventricular tachycardia. There were no sustained arrhythmias, syncope, or cardiac arrest. Our findings are similar to rates described previously in pediatric patients with CM (8, 9, 14), in which less than 1% of tests required termination for safety concerns. Patients that underwent CPETs were referred by cardiologists. Thus, there is likely a degree of preselection bias of patients deemed reasonable candidates for CPET, which may lead to artificially high exercise values and underestimation of CPET risk in the CM group compared to a global analysis of all patients with CM. Symptoms during CPETs were generally more prevalent in control patients, which is likely related to the rationale for performing CEPT given control patients were referred for symptoms with exercise or family history of cardiac disease. Control patients reported significantly more chest pain and dizziness, with no difference in palpitations, lightheadedness, or shortness of breath between CM and controls. Prior work has demonstrated VO_2_ peak does not correlate well with New York Heart Association class (18), suggesting that relying on self-reported symptoms may be an ineffective method of screening risk in CM patients. Given that there was a low prevalence of reported symptoms in our CM group, using CPET as routine surveillance to better understand CM patients’ functional capacity may be more useful than using reported symptomatology as a criterion to proceed with a CPET.

Patients with CM had impaired exercise capacity, as assessed by peak and submaximal VO_2,_ which can be a poor prognosticator in patients with HCM, DCM, and RCM (8, 9, 11, 12). Our study observed this phenomenon of decreased exercise capacity across all CM types compared to healthy controls. Maximal CPET, defined as a respiratory exchange ratio (RER) ≥ 1.0 and subjective assessment, was achieved in 100% of RCM patients, 67% of LVNC patients, 64% of HCM patients, 60% of other CM, and 50% of DCM. Due to insufficient power, we were unable to compare exercise parameters between CM subtypes. However, peak exercise values (VO_2_, SBP, HR, and VE) and submaximal VO_2_ were all lower in CM patients compared to controls. We believe that the lower peak and submaximal VO_2_ seen in CM patients is likely a reflection of the lower cardiac output in CM compared to controls, the combined effect of reduced HR and stroke volume.

Ventilatory inefficiency has previously been associated with increased risk of major events (cardiac-related death, heart transplant, and functional deterioration) in adults with HCM (19). VE/VCO_2_ slope was not different between CM patients and controls in our cohort, which was discordant from prior small studies in pediatric DCM (with heart failure) and RCM, possibly due to worse clinical status in other studies (12, 13). However, lower VE was seen in CM patients despite no significant difference in peak respiratory rate from controls. There was a higher prevalence of restrictive lung disease on baseline PFTs in CM patients versus controls, which correlated with lower VE and lower peak VO_2_, suggesting potential differences in tidal volumes with exercise in patients with CM. There is limited data on VE, exercise tidal volume, and peak respiratory rate in CM. However, a small study in children with DCM (13) reported lower VE and lower tidal volumes at peak exercise compared to controls, with no difference in respiratory rate between groups, which matches the findings from our study. The mechanism that contributes to this difference is unclear, but it may be possible that myopathy within the respiratory muscles contributes to reduced tidal volume and VE. In patients post-Fontan, targeted respiratory muscle training in conjunction with exercise training has been shown to result in increased VE at peak exercise (20). Respiratory muscle training in CM may be a future area for focused intervention to improve exercise capacity.

Abnormal blood pressure response to exercise, defined as a decrease or increase in SBP <20 mmHg compared to baseline, has been associated with increased risk of non-sudden cardiac death. Prior studies have shown the majority of HCM patients on BP lowering agents maintain a normal BP response to exercise (17). Only two patients (one on beta-blocker, one on no medication) in our cohort demonstrated relative hypotension with exercise. Peak systolic BP was significantly lower in CM patients (p= 0.021). There was no significant difference in baseline or peak BP for CM patients on or off an ACE inhibitor (p=0.333) or beta blocker (p=0.104).

Limitations of this study include selection bias of CM patients that were referred for CPET, presumably deemed reasonable candidates by their respective providers, and may have underrepresented patients with advanced disease. Additionally, we are limited by this study being a retrospective analysis. Given the small number of patients, we did not have sufficient power to do multivariate analysis between CM groups. However, this study represents one of the largest datasets for CPET data in children and adolescent CM patients, highlighting the importance of international CM data registries to include CPET data. Additionally, due to multiple missing data points, we could not properly investigate the relationship between CPET and echocardiogram, ECG, Holter, and laboratory results. Lastly, our CPETs were performed at higher elevation than most other institutions and exercise parameters (such as VO_2_) may vary with altitude (21–22).

## Conclusion

In our cohort, CPET in children and adolescents with cardiomyopathy (including HCM, DCM, RCM, LVNC, Other) is safe. Peak exercise values (VO_2_ peak, SBP, HR, VE) and submaximal VO_2_ were lower in CM patients compared to age-matched controls, likely reflective of decreased cardiac output in CM patients. These findings suggest that it is safe to perform routine CPETs to objectively monitor the impact of CM on functional status. A larger study is needed to validate our results, particularly at sea level, including assessment of whether serial CPET variables in children and adolescents with CM can be used as prognostic indicators for outcomes, including transplant and death.

## Data Availability

The authors declare that all supporting data are available within the article [and its online supplementary files].

## Acknowledgments

Kelsey Iguidbashian, MD lead project idea, data collection and writing of manuscript. Julie Fernie, MS contributed to project idea, data collection and interpretation, and writing. Kaitlin Olson, MS provided statistical analysis. Jean Cavanaugh, PA-C contributed to patient selection. Shelley Miyamoto, MD contributed to project idea and writing. Roni Jacobsen, MD contributed to project idea, interpretation and writing.

## Abbreviations

CPET: cardiopulmonary exercise testing
HCM: hypertrophic cardiomyopathy
CM: cardiomyopathy
BMI: body mass index
HR: heart rate
BSA: body surface area
ECG: electrocardiogram
VE: ventilation
RCM: restrictive cardiomyopathy
DCM: dilated cardiomyopathy
LVNC: left ventricular non-compaction
ARVC: arrhythmogenic right ventricular cardiomyopathy
PFT: pulmonary function testing
VO_2_: oxygen consumption
VCO_2_: carbon dioxide production
RER: respiratory exchange ratio
SpO_2_: oxygen saturation
BP: blood pressure
LV: left ventricular
PVC: premature ventricular contraction
ACE: angiotensin-converting enzyme

